# Multi-Omic Profiling Reveals Antibody-Drug Conjugate Targetability in Ovarian Cancer

**DOI:** 10.64898/2026.03.25.26349056

**Authors:** Elsi Pöllänen, Taru A. Muranen, Alexandra Lahtinen, Kaiyang Zhang, Daria Afenteva, Anna Pirttikoski, Susanna Holmström, Yilin Li, Kari Lavikka, Jaana Oikkonen, Jenni Söderlund, Johanna Hynninen, Anni Virtanen, Sampsa Hautaniemi

## Abstract

Antibody-drug conjugates (ADCs) require high and homogeneous target expression for optimal efficacy, yet the spatial, temporal, and cellular heterogeneity of clinically approved ADC targets in high-grade serous ovarian cancer (HGSC) remains incompletely defined. We analyzed bulk RNA-sequencing, single-cell RNA-sequencing, and whole-genome sequencing data from 867 samples across 304 patients enrolled in the real-world DECIDER cohort to systematically evaluate 11 approved ADC targets. *FOLR1, TACSTD2*, and *ERBB2* emerged as highly expressed candidates. Inter-patient variability exceeded intra-patient heterogeneity, which further decreased following neoadjuvant chemotherapy. Target expression was highly concordant across anatomical sites and largely stable from diagnosis to relapse. Single-cell RNA-sequencing results revealed that *TACSTD2* and *FOLR1* showed the most frequent cancer cell-restricted expression. In rare cases of gene amplification, *ERBB2* and *F3* emerged as potential targets alongside *TACSTD2* and *FOLR1*. Overall, 80% of patients displayed homogeneous expression of at least one actionable target, with frequent co-expression of *TACSTD2* and *FOLR1*. These findings indicate that ADC target expression in HGSC is broadly stable across space and time and support the prioritization and strategic integration of *TACSTD2*- and *FOLR1*-directed ADCs in this disease.

## Introduction

Antibody-drug conjugates (ADCs) are a class of rapidly expanding targeted therapies used in the field of precision oncology^1^. ADCs have attracted considerable attention due to their superior activity, reduced systemic exposure, and their potential to treat cancers that are resistant to traditional chemotherapy compounds^2,3^. In an ADC, a cytotoxic payload is chemically linked to a monoclonal antibody that recognizes and delivers it directly to cancer cells^4^. Following their mechanism of action, the efficacy of ADCs is reliant on high and cancer cell-specific target antigen expression, while the poorly-understood spatial and temporal heterogeneity can limit clinical activity^2,5–8^.

Dependence on the target antigen makes ADCs a suitable class of therapeutics for a histology-agnostic expansion of indication, and in ovarian high-grade serous carcinoma (HGSC), treatment based on antigen expression has shown potential^2,9^. HGSC, the most common subtype of ovarian cancer, is considered difficult-to-treat due to high heterogeneity, abundant copy number variations, and high recurrence rate accompanied by chemoresistance^10–12^. In 2022, FDA granted approval for Mirvetuximab Soravtansine, the first ADC for platinum resistant ovarian cancer, bringing onboard an alternative treatment option for patients with HGSC^13,14^. Since then, multiple ADCs have shown antitumor activity in clinical trials, highlighting the potential of the class of therapeutics in ovarian cancer^5^.

Herein, we used real-world patient data from the DECIDER clinical trial (NCT04846933) to characterize the expression profiles of genes encoding approved ADC targets in HGSC and to identify patient subgroups most likely to benefit from treatment. We hypothesize that approved ADCs can be repurposed for HGSC when their target antigens exhibit high and homogeneous expression.

## Materials and methods

### DECIDER participants

In this study, we used bulk RNA-sequencing (RNA-seq), single-cell RNA-sequencing (scRNA-seq), and whole-genome sequencing data from the ongoing, observational DECIDER clinical trial (NCT04846933). Patients were recruited between February 2010 and March 2024, and treated at Turku University Hospital, Finland. Primary therapy comprised 163 patients treated with primary debulking surgery (PDS) followed by median of six cycles of platinum-taxane chemotherapy and 141 patients treated with median of three cycles of neo-adjuvant chemotherapy (NACT) followed by interval debulking surgery (IDS) and median of three cycles of adjuvant chemotherapy. The study was approved by the Ethics Committee of the Hospital District of Southwest Finland (VARHA/28314/13.02.02/2023) and is being conducted in accordance with the ethical principles outlined in the Declaration of Helsinki. All patients provided written informed consent.

### Sample selection

Samples were collected from several anatomical sites including primary tubo-ovarian (adnex) sites (N = 246), intra-abdominal solid metastases (N = 480), distant metastases (N = 31), including lymph nodes, pleural fluid and vagina, and from ascites (N = 110). Untreated diagnostic samples (N = 657) were collected during diagnostic laparoscopy (NACT-patients) or PDS (PDS-patients), post-NACT samples (N =154) during interval debulking surgery (only NACT-patients), and relapse samples (N = 56) at cancer progression. Valid sequencing data was required to have an estimated tumor cell fraction of at least 0.3 from the PRISM^15^ estimation and at least 0.1 from the ASCAT^16^ v2.5.2 estimation.

### Bulk RNA-sequencing data

Quality controlled bulk RNA-seq data was available for 867 samples and 304 patients. Bulk RNA-seq data was processed as previously described^17^. To account for the varying cancer compositions in the surgically collected patient samples, we decomposed the bulk RNA-seq signal using PRISM^15^ v0.9 and all analyses were run in the cancer cell specific component. The decomposed bulk RNA-seq data was TPM-normalized using one representative transcript for each gene. Matched annotation between NCBI and EMBL-EBI (MANE) v1.0 transcript was used when available for the gene in GENCODE v25 annotation^18^.

### Single cell RNA-sequencing data

scRNA-seq data was available for 95 samples and 57 patients and processed as previously described^19,20^.

### Whole-genome sequencing data

Matching whole-genome sequencing data was processed as previously described^20^. The copy-number variation analysis was computed as previously described^21^. Briefly, we ran CATK^22^ v4.1.4.1 for copy-number segmentation and reimplemented ASCAT^16^ algorithm to estimate purity, ploidy and purified logR values.

### Mean expression distribution

We computed mean expression values of all protein-coding genes throughout the cohort. The mean expression distribution was divided into quartiles, and the threshold value of fourth quartile marked the limit of high expression. The threshold for detectable expression was set to median, with a rough assumption that approximately 50% of all protein-coding genes are not expressed in cells. Analysis was run using an R environment for statistical computing v4.2.0^23^.

### Intra- and inter-patient heterogeneity

We used a linear mixed effects model to study intra- and inter-patient heterogeneity in expression as previously described^24^. The model was in parallel fitted to sample-subgroups from diagnosis and IDS using R package lme4 v1.1.35.5. The goodness of fit was reported for fixed effects (R-squared marginal, R2m) and for the complete model (R-squared conditional, R2c) using R package MuMin 1.48.4.

Briefly, inter-patient heterogeneity was defined as the proportion of variance explained by the random effect for patient, while intra-patient heterogeneity accounted for the variance rising from systematic tumor site differences combined with the variance explained by patient-specific site deviations. Using R package emmeans v1.10.6, we derived the estimated marginal means for the two tumor site groups studied here: adnex and metastases (solid intra-abdominal metases and lymph nodes). Tukey honest significance difference adjustment was used to control for the multiple pairwise comparisons and results were considered statistically significant when the adjusted p-value was < 0.05.

We analyzed the target expression in a subset of patients with diagnostic samples from adnex and the metastatic sites included in the model, to further assess the significance of systematic variation. For each of the analyzed targets, the patient-matched sample pairs of adnex and metastasis were illustrated in a scatterplot and divided into four sections based on the target-specific estimated marginal mean for adnex. Percentages of data points belonging to each section were reported on the plot. If a patient had several available samples, pairing was performed exhaustively, such that all available adnex samples were matched with all available metastasis samples.

To understand the effect that treatment might have on the target expression, we performed patient-matched exhaustive pairing of diagnostic samples with samples collected from IDS and relapse. The representative sample pair for each patient was the pair with the highest EOC purity estimate, and matching tumor sites and solid tissues were prioritized in the selection. Statistical significance of the target expression change was confirmed using paired t-test and the results were considered statistically significant when the adjusted p-value was < 0.05. All heterogeneity analyses were run in R software v4.4.2.

### Intra-tumor heterogeneity

The intra-tumor heterogeneity of target gene expression was assessed using scRNA-seq data. Samples containing less than 40 cancer cells were excluded from the downstream analysis, as the same cell number cut-off has previously been used as the inclusion criteria for pseudobulk analysis^25^.

Similarly to the bulk RNA sequencing data, the detectable expression threshold was derived using mean expression distribution for the matching set of protein coding genes and the cancer cell pool from the Seurat object. Means were computed across all available cancer cells instead of samples. For each sample, individual cancer cells were classified to be positive if their target expression exceeded the detectable expression median. Samples with at least 75% positive cells were considered homogeneously expressing the target. To define RNA-level criteria for classifying eligible patients for treatment with various ADCs, we aggregated the numbers of homogeneously expressing samples to patient level so that a patient was classified as positive for a target if all their samples exceeded the threshold of 75% of the cells expressing the given target.

To further annotate the cell-specific expression levels, we introduced three novel categories to the used expression distribution. The three categories corresponded to the highest 10%, highest 1%, and the highest 0.1% of mean expression distribution. All cells within a sample were then annotated in one of the five possible categories and plotted together with the number of cells within the sample to demonstrate deeper expression annotation for all analyzed samples.

Finally, the relationship between *FOLR1* and *TACSTD2* expression was assessed by integrating intratumor analyses with the bulk RNA-seq data. The optimal threshold for the bulk RNA-seq values were determined using receiver operating characteristic (ROC) curve analysis, selecting the cut-off that maximized Youden’s index. Analysis was conducted using the R package pROC v1.18.5.

### Results

To characterize the expression of 11 targets corresponding to 13 clinically approved ADCs (Table 1), we analyzed gene expression data from 867 samples and 304 patients with HGSC enrolled in the DECIDER trial, complemented by single-cell RNA sequencing and whole-genome sequencing datasets. To enable relative expression stratification, gene expression levels were classified into quartiles using cohort-wide expression means of all protein-coding genes (Figure 1A). ADC targets were stratified into three tiers (Figure 1B): Tier 1, mean expression in Q4 (*FOLR1, TACSD2, ERRB2*); Tier 2, mean expression in Q2–Q3 with at least one sample in Q4 (*F3, NECTIN4, EGFR, CD22*); and Tier 3, mean expression in Q1 (*TNFRSF8, CD19, CD33, CD79B*). As expected, almost all ADC targets for hematological malignancies fell to into Tier 3 and the elevated expression of the B-cell marker *CD22* appeared to be explained by recurrent amplification of the centromeric region on chromosome 19. Due to the low expression of Tier 3 targets throughout the whole cohort, we excluded them from subsequent analyses.

**Table 1.** Approved ADCs. The list and characteristics of globally approved and commercially available ADCs by the end of 2024^26^.

| Antibody-drug conjugate | Target antigen | Accepted | Original indication |
| --- | --- | --- | --- |
| Mirvetuximab soravtansine | FOLR1 | U.S. FDA (Nov, 2022) | FRα-positive epithelial ovarian cancer |
| Sacituzumab govitecan | TACSTD2 | U.S. FDA (Apr, 2020) | Metastatic triple-negative breast cancer |
| Trastuzumab deruxtecan | ERBB2 | U.S. FDA (Dec, 2019) | Unresectable HER2-positive breast cancer |
| Disitamab vedotin | ERBB2 | China NMPA (Jun, 2021) | Advanced or metastatic gastric cancer |
| Trastuzumab emtansine | ERBB2 | U.S. FDA (Feb, 2013) | HER2-positive metastatic breast cancer |
| Enfortumab vedotin | NECTIN4 | U.S. FDA (Dec, 2019) | Advanced or metastatic urothelial cancer |
| Tisotumab vedotin | F3 | U.S. FDA (Sep, 2021) | Recurrent or metastatic cervical cancer |
| Cetuximab sarotalocan | EGFR | Japan PMDA (Sep, 2020) | Advanced or recurrent head and neck cancer |
| Loncastuximab tesirine | CD19 | U.S. FDA (Apr, 2021) | Relapsed or refractory large B-cell lymphoma |
| Polatuzumab vedotin | CD79B | U.S. FDA (Jun, 2019) | Diffuse large B-cell lymphoma |
| Inotuzumab ozogamicin | CD22 | U.S. FDA (Aug, 2017) | B-cell precursor acute lymphoblastic leukemia |
| Brentuximab vedotin | TNFRSF8 | U.S. FDA (Aug, 2011) | Hodgkin lymphoma; Large-cell lymphoma |
| Gemtuzumab ozogamicin | CD33 | U.S. FDA (Mar, 2000) | CD33-positive acute myeloid leukemia |

**Figure 1.**
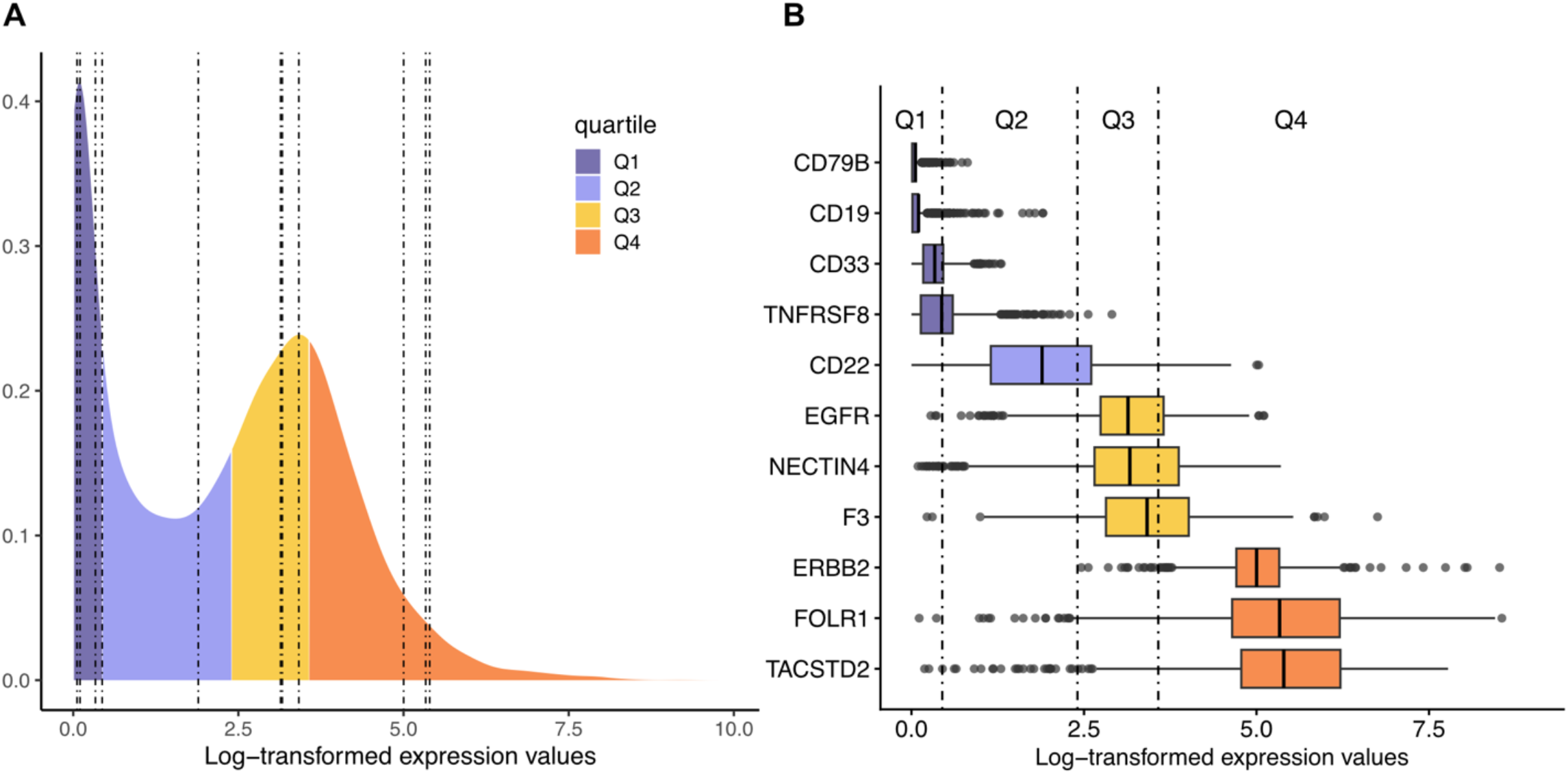
Expression distribution of the ADC targets. **A**. The global mean expression landscape of the protein coding genes in HCSC. The quartiles are marked in the illustration with different colors, and the ADC targets with vertical dashed lines. **B**. Target-specific expression profiles as boxplots. ADC-targets are colored based on the quartile ranges. Means marked in the boxplots match the vertical lines in figure 1A and the vertical lines mark the thresholds of the different quartiles.

### Treatment enhances intra-patient homogeneity of Tier 1 and Tier 2 targets

To quantify intra- and interpatient heterogeneity in Tier 1 and Tier 2 targets, we applied a multivariable random-effects model (Methods). We first confirmed that the model fit the data well for most targets, both before and after neoadjuvant chemotherapy (NACT, Supplementary Material). Briefly, at diagnosis the fit was 66.0% on average (range: 0.524-0.775) and after NACT 65.5% (range: 0.419-0.816). Interestingly, inter-patient heterogeneity increased, and intra-patient heterogeneity decreased for most ADC targets after NACT (Supplementary Table 1).

Our results show that tumor site accounted for only a minor fraction of the observed variance, with a further reduction following NACT (Figure 2A). Some statistically significant differences were detected at the cohort level (Supplementary Figure 1); however, the patient-matched samples did not demonstrate a systemic effect (Figure 2B). Moreover, results indicate that intra-patient heterogeneity was primarily driven by low-expression samples (Figure 2B). Collectively, these data support the generalizability of tissue sampling for Tier 1 and Tier 2 targets.

**Figure 2.**
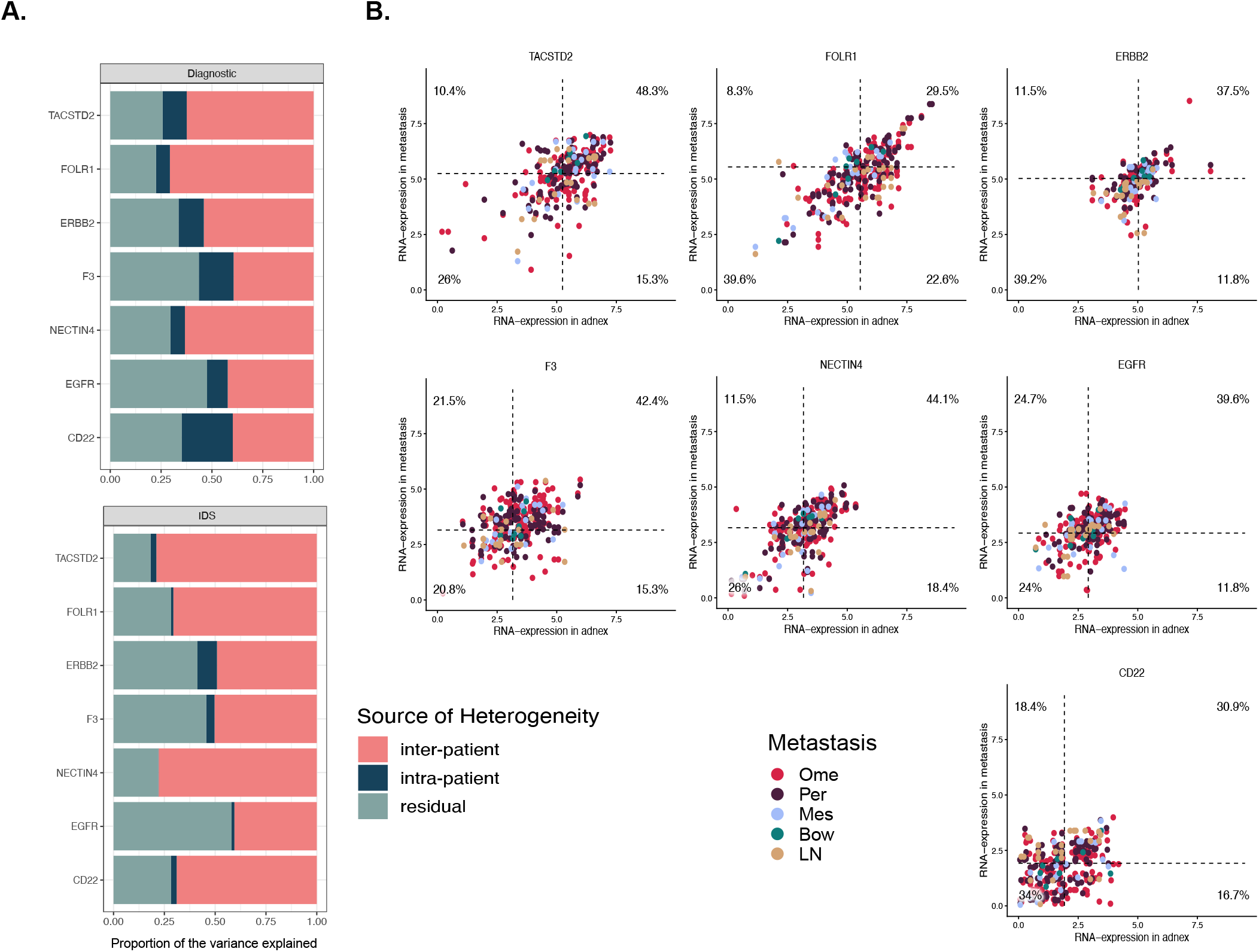
Source of heterogeneity. **A.** The proportions of variance explained by intra-patient, inter-patient, and residual heterogeneity, which reflects the unexplained portion of the total variance. Proportions are reported for diagnostic and IDS samples separately. **B**. The scatterplot of the patient matched pairs of adnex and metastatic samples. Specific metastatic sites are marked with different colors. Target specific EMMs for adnex are used to divide the observations into four groups. The percentage of observations belonging to each group are marked in the four corners of the illustrations.

### Target expression remains comparable from diagnosis to relapse

To assess the temporal dynamics of target expression, we analyzed matched patient sample pairs collected at diagnosis and after NACT at interval debulking surgery (IDS), as well as matched samples obtained at diagnosis and relapse. None of the targets exhibited a statistically significant systemic decrease in expression in either longitudinal analysis. However, we observed substantial variability in individual cases (Figure 3 and Supplementary Figure 2). Notably, for *TACSTD2* and *FOLR1*, individual cases with high expression in diagnosis showed drastic decrease in expression in the matched IDS sample (Supplementary Figure 2).

**Figure 3.**
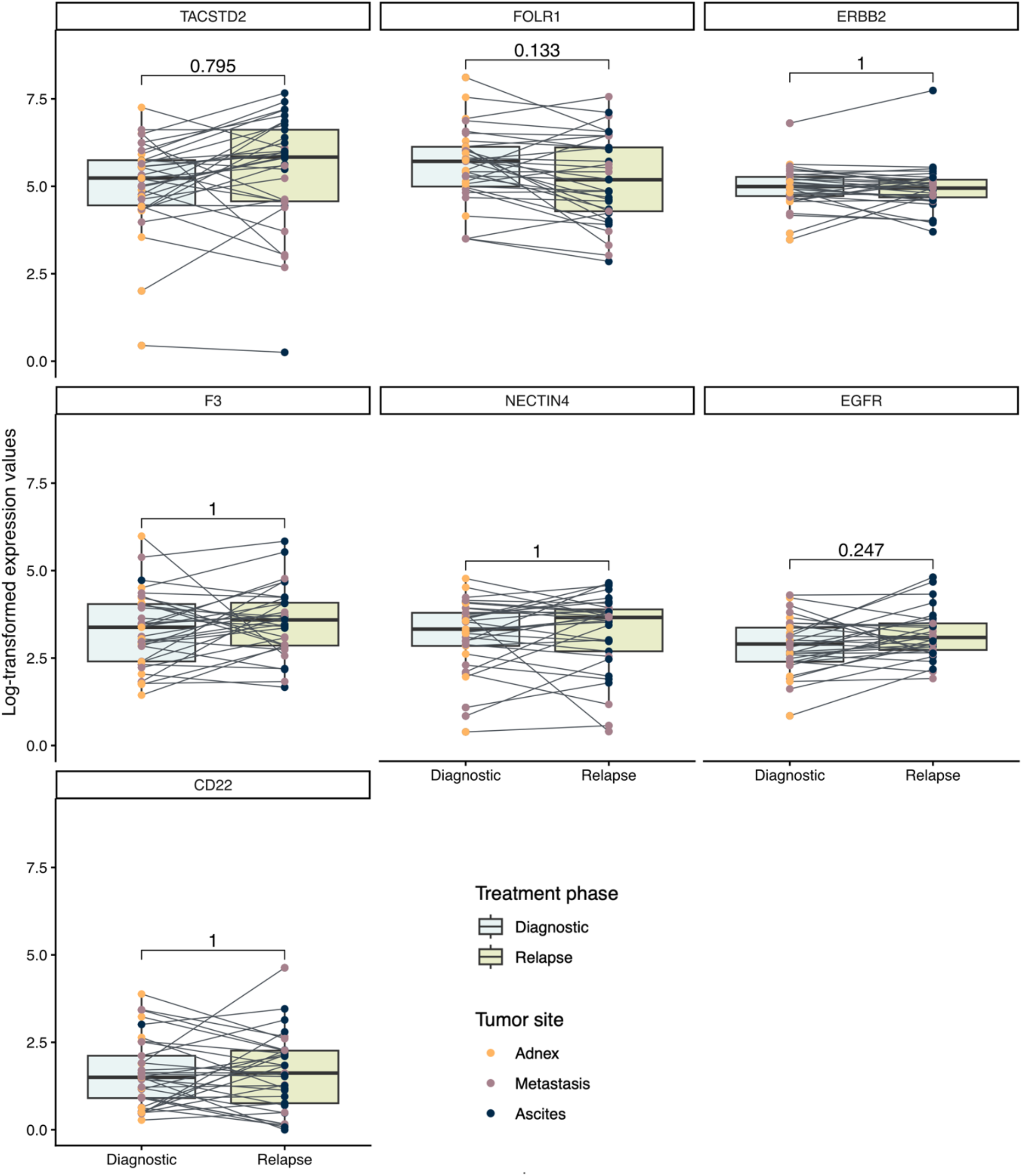
Target expression across diagno and relapse. The figure illustratesthe RNA expression of patient matched diagnostic and relapse samples. Each smpie is coiored on the collected tumor site, and each diagnostic-relapse pair relates to a line.

In matched diagnosis–relapse pairs, we did not observe significant changes in target expression across ADC targets (Figure 3). Importantly, this suggests that, despite variable treatment exposures and time intervals between diagnosis and relapse, target expression at diagnosis remains informative for the relapse setting. Although individual cases with variable expression across timepoints were observed, the differences were less pronounced than those observed for diagnosis–IDS pairs. This might have been partially explained by the absence of relapse sample for the extreme *TACSTD2* and *FOLR1* cases identified in the diagnosis–IDS pairs. *TACSTD2* was the only target showing a trend toward increased expression at relapse, an effect that was enriched in samples derived from ascites.

### Intra-tumor heterogeneity analysis reveals competing ADC targets

Single-cell RNA sequencing of 95 HGSC samples was used to evaluate intratumoral heterogeneity, with UMAP visualization shown in Figure 4A. *FOLR1, NECTIN4*, and *ERBB2* demonstrated cancer cell-restricted expression. *TACSTD2* was highly expressed in cancer cells but also showed low-level expression across other cell types, including immune cells. *EGFR* and *F3* were broadly expressed across malignant and non-malignant populations, whereas *CD22* expression was lower in cancer cells than in other cell types, such as B cells and mast cells.

**Figure 4.**
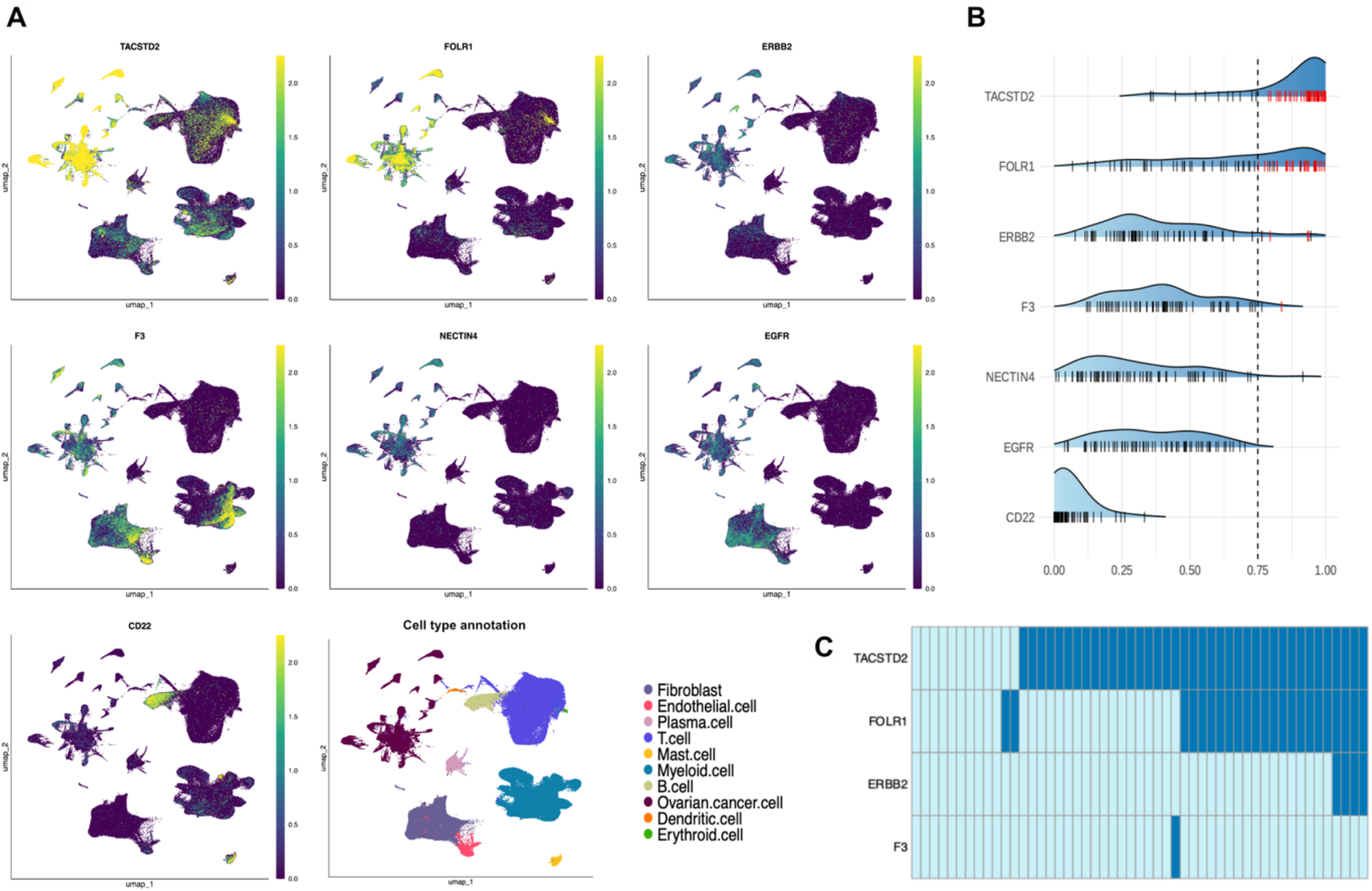
Intra-tumor heterogeneity. **A**. Feature plots illustrating ADC-target gene expression across the different cell types. **B**. Intra-tumor heterogeneity is illustrated as the percentage of cells highly expressing target genes. Each sample is marked as a black line if the sample is from a patient that is negative for the given target or red is the patient is positive. The threshold 0.75 is marked with the vertical dashed line. **C**. Heatmap presenting the ADC targets with a potential target cohort. Positive cases are marked with dark blue and negative cases with light blue.

To quantify intratumoral heterogeneity, we calculated, for each sample, the proportion of cancer cells expressing each target above the detection threshold (Figure 4B). Samples were classified as positive if more than 75% of cancer cells exceeded this threshold (Methods), and patients with exclusively positive samples are shown in Figure 4C. Importantly, 41/51 patients were positive for at least one target. *TACSTD2* showed the highest number of patients with homogeneous expression (*n* = 39, 76%), followed by *FOLR1* (*n* = 23, 45%). In contrast, *ERBB2* and *F3* were homogeneously expressed in only a small number of cases (*n* = 4 for *ERRB2* and *n* = 1 for *F3*).

We detected strong co-expression of targets with the greatest overlap observed between *TACSTD2* and *FOLR1* (Figure 4C). Nearly all *FOLR1*-positive patients also showed homogeneous *TACSTD2* expression. Neither *ERBB2* nor *F3* occurred as a stand-alone target; all *ERBB2*-positive patients co-expressed *TACSTD2* and *FOLR1*, and the single *F3*-positive case was also *TACSTD2*-positive. Unlike *TACSTD2* and *FOLR1*, homogeneous expression of *ERBB2* and *F3* was amplification-driven (50% for *ERRB2* and 100% for *F3*).

Intratumor heterogeneity estimates were generally concordant with bulk RNA-seq expression levels (Supplementary Figure 3). *FOLR1* and *TACSTD2* remained homogeneously expressed across different bulk RNA-seq thresholds, showing strong overlap in expression patterns, though complementary cases were also identified. (Supplementary Figure 4). While the proportion of cells above the detection threshold was lower for *FOLR1*, deeper annotation of the cell-specific expression levels (Methods) revealed high expression of *FOLR1* in at least half of the cells in most samples (Figure 5). Also *ERBB2, F3, NECTIN4*, and *EGFR*, reached the highest 90% of the expression in most samples. However, these targets were highly expressed in considerably smaller subsets of cells, with the proportions rarely reaching 0.5 (Figure 5). Collectively, these data support the preferential selection of *TACSTD2*- and *FOLR1*-targeted ADCs in HGSC.

**Figure 5.**
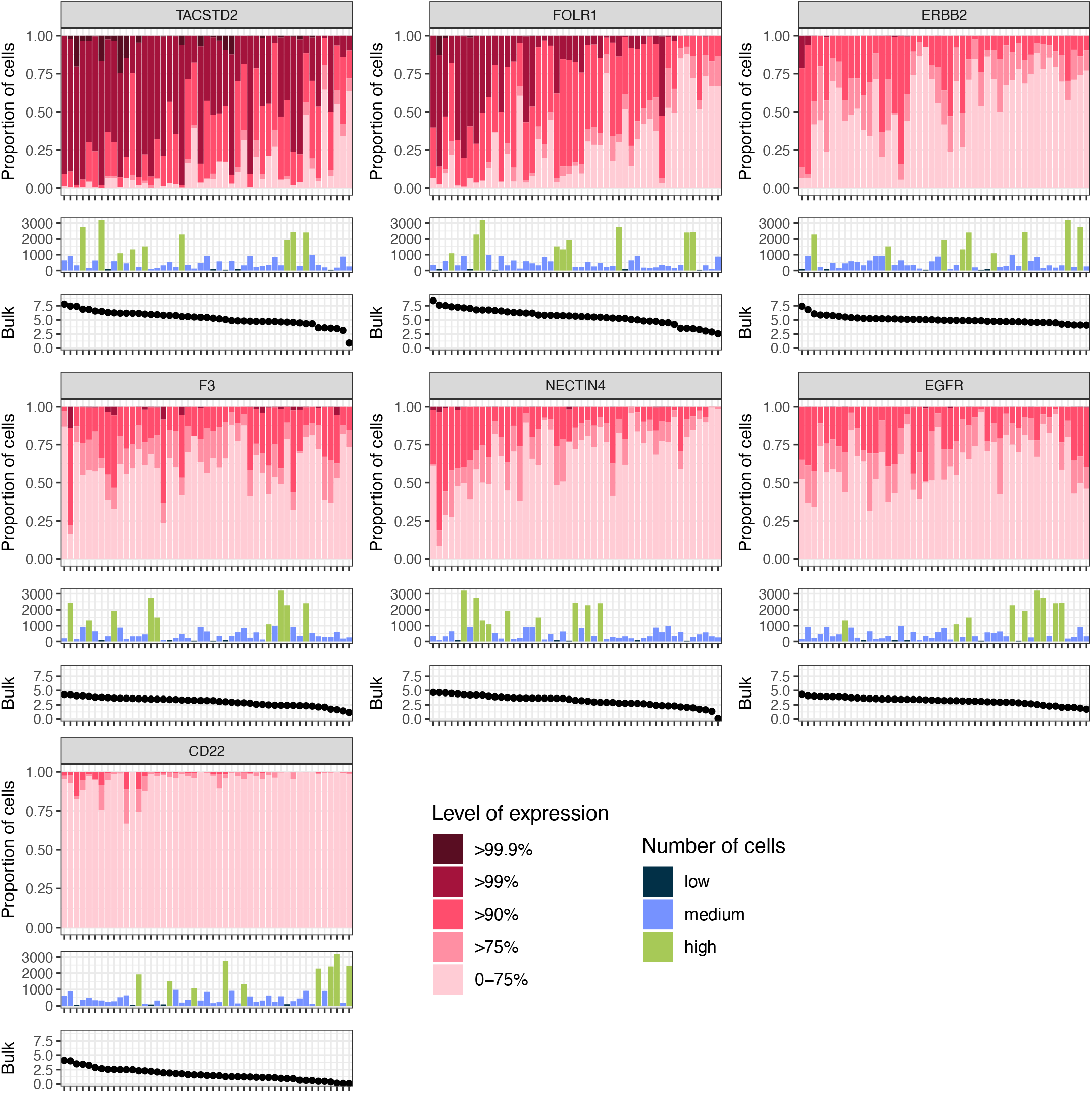
Cell-specific expression annotation. Deeper cell-specific expression annotation for the single-cell samples with available matched bulk RNA-seq data. The first plot represents the proportion of cancer cells and their expression levels distinguished by colors. The second plot shows the number of cancer cells in for those samples, and the third plot shows the expression in the matched bulk RNA-seq data. Each target-specific panel of the three plots is ordered based on decreasing expression of the bulk RNA-seq data.

## Discussion

To answer the need for treatment options for patients with relapsed HGSC, we characterized the potential of clinically approved 13 ADCs^26^ based on their targets’ gene expression in the real-life DECIDER observational trial. We analyzed expression levels and heterogeneity of the ADC target genes across 304 patients with HGSC. In addition to *FOLR1*, which is already approved for HGSC, our results support the utility of using ADCs targeting *TACSTD2, ERBB2*, or *F3* in HGSC.

Heterogeneity is a key challenge of the clinical efficacy of ADCs^5,27^. As expected, the level of inter-patient heterogeneity surpassed intra-patient heterogeneity. Earlier studies have hypothesized that varying tumor settings, such as physical site or treatment phase, affect the clinical activity of ADCs ^28,29^. Our results show that target expression patterns were highly concordant between primary adnexal tumors and matched metastatic lesions, indicating that sampling from either site provides a reliable representation of patient-specific target expression levels. Furthermore, diagnostic samples can be used to approximate the target expression during IDS and relapse, suggesting that diagnostic samples are amenable for ADC testing.

Using single-cell RNA sequencing from 95 patients, we systematically quantified intratumoral heterogeneity and found that ADC target expressions exhibit widely divergent heterogeneity profiles within tumors, underscoring fundamental differences in their cellular distribution. Both *FOLR1* and *TACSTD2* are homogeneously expressed in the tumor cells, which supports their utility in HGSC as observed previously ^14,30,31^. Notably, *F3* and *ERBB2* exhibited homogeneous expression in tumors harboring amplification of their respective gene loci, consistent with copy-number gain as an early tumorigenic event that becomes ubiquitously represented across tumor subclones. The clonal stability of amplification-driven targets may limit phenotypic evasion, thereby posing a prioritization challenge when selecting among multiple ADC-eligible targets.

We found multiple ADC targets simultaneously actionable within individual patients, underscoring both the need for rational prioritization strategies to guide ADC selection and the opportunity for combinatorial therapeutic approaches. Our data point to novel therapeutic opportunities involving the pairing of anti-*TACSTD2* and anti-*FOLR1* ADCs. Sequential treatment strategies alternating distinct ADCs have previously demonstrated clinical benefit^32–34^, where the antitumor activity was proposed to arise from the use of mechanistically distinct payloads exemplified by mirvetuximab soravtansine and sacituzumab govitecan^2,33^.

Novel ADC incorporating a bispecific antibody targeting *TACSTD2* and *FOLR1* could be considered for the dual-expressing tumors. Previously dual-targeting strategies have been associated with enhanced tumor specificity^35^ and, in some settings, superior efficacy compared to single-target ADCs^36^. Notably, although the safety profiles of the ADCs examined here have been individually characterized, the broad background expression of *TACSTD2* suggests that bispecific ADC designs may mitigate potential on-target, off-tumor toxicities by increasing tumor-selective engagement.

In summary, we have performed systematic and comprehensive characterization of ADC targets’ gene expression profiles before and after chemotherapy in matched samples and tissues. Our results show that HGSC, cancer that is challenging to manage, is amenable for multiple ADC targets as only 20% of the patients were negative for all studied targets. Thus, ADCs are a promising avenue to improve survival of patients with HGSC.

## Limitations of the study

We acknowledge that successful treatment with ADCs is dependent on the protein expression of the target gene, and the missing information of protein existence to be one of the limitations of our study. However, positive correlation between ADC-targets’ mRNA and protein expression has previously been shown^37^, which supports using transcriptomics data in building reliable target gene expression profiles. Additionally, our approximated 45% of potential patient cohort benefitting from *FOLR1* targeted therapy was very close to the previously reported 36% of patients eligible to treatment with Mirvetuximab Sorvatansine in the SORAYA study^38^. For those ADCs that are known to exert their cytotoxic activities through bystander effect^39^, the estimated target cohort might be bigger since lower target expression could be considered^34^.

## Data Availability

Data availability: Raw WGS, bulk RNA-seq and scRNA-seq data has been deposited at the European Genome-phenome Archive (EGA), which is hosted by the EBI and the CRG, under study accession numbers EGAS00001006775, EGAS00001004714 and EGAS00001005010, respectively.

## Acknowledgements

The authors are grateful to the patients for their participation, the Turku University Hospital staff for recruitement and sampling, and all research nurses, technical assistance, and administrative personnel, especially Peppi Alho, Elina Valkonen, Nina Halme, Ann-Christin Ostwaldt, and Jenni Lahtinen. The authors wish to acknowledge the CSC - IT Center for Science (Finland) for computational resources. Funding: This project has received funding from the European Union’s Horizon 2020 Research and Innovation Programme under grant agreement 965193 for DECIDER; the Sigrid Jusélius Foundation; and the iCANDOC Presicion Cancer Medicine pilot. The funders had no role in the design of the study; in collection, analysis and interpretation of data; or in preparing the manuscript.

Authors contributions: Conceptualization: T.A.M., A.L., E.P., A.V., J.H., J.S, and S.H.; Methodology and Software: E.P., K.Z., D.A., and A.P.; Formal Analysis: E.P., T.A.M, and K.Z.; Investigation: E.P., T.A.M., A.L., A.V., J.H., J.S., and S.H.; Resources: E.P., A.L., D.A., Su.H., Y.L., K.L., J.O., J.S., J.H., and S.H.; Data curation: D.A., A.P., Su.H., Y.L., K.L., J.O., J.S., J.H., A.V.; Visualization: E.P.; Supervision: T.A.M., A.L., and S.H.; Project Administration: J.H., A.V., and S.H.; Funding Acquisition: E.P., and S.H.; Writing Original Draft: E.P.; Writing-Review and Editing: All authors reviewed and approved the manuscript. Competing interests: The authors declare no competing interests. Data availability: Raw WGS, bulk RNA-seq and scRNA-seq data has been deposited at the European Genome-phenome Archive (EGA), which is hosted by the EBI and the CRG, under study accession numbers EGAS00001006775, EGAS00001004714 and EGAS00001005010, respectively.

## References

1. Colombo, R., Tarantino, P., Rich, J. R., LoRusso, P. M. & de Vries, E. G. E. The Journey of Antibody–Drug Conjugates: Lessons Learned from 40 Years of Development. Cancer Discov. 14, 2089–2108 (2024).

2. Tarantino, P. et al. Antibody–drug conjugates: Smart chemotherapy delivery across tumor histologies. CA Cancer J. Clin. 72, (2022).

3. Moore, J. M., Bell, E. L., Hughes, R. O. & Garfield, A. S. ABC transporters: human disease and pharmacotherapeutic potential. Trends in Molecular Medicine vol. 29 Preprint at 10.1016/j.molmed.2022.11.001 (2023).

4. Tsuchikama, K., Anami, Y., Ha, S. Y. Y. & Yamazaki, C. M. Exploring the next generation of antibody–drug conjugates. Nature Reviews Clinical Oncology vol. 21 Preprint at 10.1038/s41571-023-00850-2 (2024).

5. Silverstein, J., Karlan, B., Herrington, N. & Konecny, G. Antibody-drug conjugates as targeted therapy for treating gynecologic cancers: update 2025. Current Opinion in Obstetrics and Gynecology vol. 37 Preprint at 10.1097/GCO.0000000000001002 (2025).

6. Armstrong, G. B. et al. Antibody-drug conjugates as multimodal therapies against hard-to-treat cancers. Adv. Drug Deliv. Rev. 224, 115648 (2025).

7. Loganzo, F., Sung, M. & Gerber, H. P. Mechanisms of resistance to antibody-drug conjugates. Molecular Cancer Therapeutics vol. 15 Preprint at 10.1158/1535-7163.MCT-16-0408 (2016).

8. Sharma, S., Li, Z., Bussing, D. & Shah, D. K. Evaluation of quantitative relationship between target expression and antibody-drug conjugate exposure inside cancer cells. Drug Metabolism and Disposition 48, (2020).

9. Dinkins, K. et al. Targeted therapy in high grade serous ovarian Cancer: A literature review. Gynecol. Oncol. Rep. 54, 101450 (2024).

10. Veneziani, A. C. et al. Heterogeneity and treatment landscape of ovarian carcinoma. Nature Reviews Clinical Oncology vol. 20 Preprint at 10.1038/s41571-023-00819-1 (2023).

11. Saleh, A. & Perets, R. Mutated p53 in hgsc—from a common mutation to a target for therapy. Cancers vol. 13 Preprint at 10.3390/cancers13143465 (2021).

12. Richardson, D. L., Eskander, R. N. & O’Malley, D. M. Advances in Ovarian Cancer Care and Unmet Treatment Needs for Patients with Platinum Resistance: A Narrative Review. JAMA Oncology vol. 9 Preprint at 10.1001/jamaoncol.2023.0197 (2023).

13. Dilawari, A. et al. FDA Approval Summary: Mirvetuximab Soravtansine-Gynx for FRα-Positive, Platinum-Resistant Ovarian Cancer. Clinical Cancer Research 29, 3835–3840 (2023).

14. Moore, K. N. et al. Mirvetuximab Soravtansine in FRα-Positive, Platinum-Resistant Ovarian Cancer. New England Journal of Medicine 389, (2023).

15. Häkkinen, A. et al. PRISM: recovering cell-type-specific expression profiles from individual composite RNA-seq samples. Bioinformatics 37, (2021).

16. Van Loo, P. et al. Allele-specific copy number analysis of tumors. Proc. Natl. Acad. Sci. U. S. A. 107, (2010).

17. Afenteva, D. et al. Multi-Omics Analysis Reveals the Attenuation of the Interferon Pathway as a Driver of Chemo-Refractory Ovarian Cancer. bioRxiv 2024.03.28.587131 (2024) doi:10.1101/2024.03.28.587131.

18. Morales, J. et al. A joint NCBI and EMBL-EBI transcript set for clinical genomics and research. Nature 604, (2022).

19. Pirttikoski, A. et al. Conserved cell state dynamics reveal targetable resistance patterns in ovarian high-grade serous carcinoma. Preprint at 10.1101/2025.06.13.659489 (2025).

20. Micoli, G. et al. Decoding the Genomic and Functional Landscape of Emerging Subtypes in Ovarian Cancer. Cancer Discov. 15, 2262–2277 (2025).

21. Lavikka, K. et al. Deciphering cancer genomes with GenomeSpy: a grammar-based visualization toolkit. Gigascience 13, (2024).

22. McKenna, A. et al. The genome analysis toolkit: A MapReduce framework for analyzing next-generation DNA sequencing data. Genome Res. 20, (2010).

23. Team, R. C. R Core Team 2023 R: A language and environment for statistical computing. R foundation for statistical computing. https://www.R-project.org/.R Foundation for Statistical Computing (2023).

24. Zhang, K. et al. Multi-modal characterization of transcriptional programs that drive metastatic cascades to solid sites and ascites in ovarian cancer. bioRxiv 2025.08.26.672372 (2025) doi:10.1101/2025.08.26.672372.

25. Perez-Villatoro, F. et al. Single-cell spatial atlas of high-grade serous ovarian cancer uncovers MHC class II as a key predictor of spatial tumor ecosystems and clinical outcomes. Cancer Discov. http://doi.org/10.1158/2159-8290.CD-25-1492 (2026) doi:10.1158/2159-8290.CD-25-1492.

26. Wang, R. et al. Antibody–Drug Conjugates (ADCs): current and future biopharmaceuticals. J. Hematol. Oncol. 18, 51 (2025).

27. Narayana, R. V. L. & Gupta, R. Exploring the therapeutic use and outcome of antibody-drug conjugates in ovarian cancer treatment. Oncogene 44, 2343–2356 (2025).

28. Ascione, L. et al. Unlocking the Potential: Biomarkers of Response to Antibody-Drug Conjugates. American Society of Clinical Oncology Educational Book 44, (2024).

29. Li, X. et al. Spatial, Temporal, and Molecular Heterogeneity of ADC targets in High-Grade Serous Ovarian Carcinoma. Preprint at 10.64898/2025.12.05.25341695 (2025).

30. Moore, K. N. et al. Phase III, randomized trial of mirvetuximab soravtansine versus chemotherapy in patients with platinum-resistant ovarian cancer: primary analysis of FORWARD I. Annals of Oncology 32, (2021).

31. Greenman, M., Bellone, S., Demirkiran, C., Max Philipp Hartwich, T. & Santin, A. D. Sacituzumab govitecan in heavily pretreated, platinum-resistant high grade serous ovarian cancer. Gynecol. Oncol. Rep. 54, (2024).

32. Banerji, U. et al. Trastuzumab duocarmazine in locally advanced and metastatic solid tumours and HER2-expressing breast cancer: a phase 1 dose-escalation and dose-expansion study. Lancet Oncol. 20, (2019).

33. Peng, X. et al. The ideal strategies of antibody‒drug conjugate sequential treatment in HER2-expressing metastatic breast cancer: A multi-center real-world study. Breast 81, (2025).

34. Modi, S. et al. Trastuzumab Deruxtecan in Previously Treated HER2-Positive Breast Cancer. New England Journal of Medicine 382, (2020).

35. Seo, H. et al. Bispecific Antibody and Antibody-Drug Conjugate as Novel Candidates for Treating Pancreatic Ductal Adenocarcinoma. Biomolecules vol. 15 Preprint at 10.3390/biom15101477 (2025).

36. Liu, C. et al. A bispecific antibody–drug conjugate targeting EGFR and HER3 in metastatic esophageal squamous cell carcinoma: a phase 1b trial. Nat. Med. 31, (2025).

37. Bosi, C. et al. Pan-cancer analysis of antibody-drug conjugate targets and putative predictors of treatment response. Eur. J. Cancer 195, (2023).

38. Matulonis, U. A. et al. Efficacy and Safety of Mirvetuximab Soravtansine in Patients with Platinum-Resistant Ovarian Cancer with High Folate Receptor Alpha Expression: Results from the SORAYA Study. Journal of Clinical Oncology 41, (2023).

39. Wang, Y. et al. Bystander effect in antibody-drug conjugates: Navigating the fine line in tumor heterogeneity. Critical Reviews in Oncology/Hematology vol. 216 Preprint at 10.1016/j.critrevonc.2025.104979 (2025).

