## Supplementary Figures for "Multi-Omic Profiling Reveals Antibody-Drug Conjugate Targetability in Ovarian Cancer"

**Supplementary Table 1. Model fit and heterogeneity proportions.**

| | R2m | R2c | Inter-patient heterogeneity | Intra-patient heterogeneity | Residual heterogeneity | Sample subset |
|---|---|---|---|---|---|---|
| TACSTD2 | 0.001 | 0.742 | 0.624 | 0.119 | 0.258 | Diagnostic |
| FOLR1 | 0.009 | 0.775 | 0.707 | 0.068 | 0.225 | Diagnostic |
| ERBB2 | 0 | 0.664 | 0.54 | 0.124 | 0.336 | Diagnostic |
| F3 | 0.011 | 0.564 | 0.394 | 0.171 | 0.436 | Diagnostic |
| NECTIN4 | 0.001 | 0.704 | 0.633 | 0.071 | 0.296 | Diagnostic |
| EGFR | 0.012 | 0.524 | 0.422 | 0.102 | 0.476 | Diagnostic |
| CD22 | 0.003 | 0.649 | 0.398 | 0.251 | 0.351 | Diagnostic |
| TACSTD2 | 0.008 | 0.818 | 0.789 | 0.028 | 0.182 | IDS |
| FOLR1 | 0.011 | 0.717 | 0.706 | 0.011 | 0.283 | IDS |
| ERBB2 | 0.005 | 0.587 | 0.491 | 0.096 | 0.413 | IDS |
| F3 | 0.041 | 0.544 | 0.503 | 0.041 | 0.456 | IDS |
| NECTIN4 | 0.001 | 0.779 | 0.778 | 0.001 | 0.221 | IDS |
| EGFR | 0.014 | 0.419 | 0.405 | 0.014 | 0.581 | IDS |
| CD22 | 0.02 | 0.718 | 0.69 | 0.028 | 0.282 | IDS |

´

### Supplementary Figure 1. Effects of the varying tumor sites.

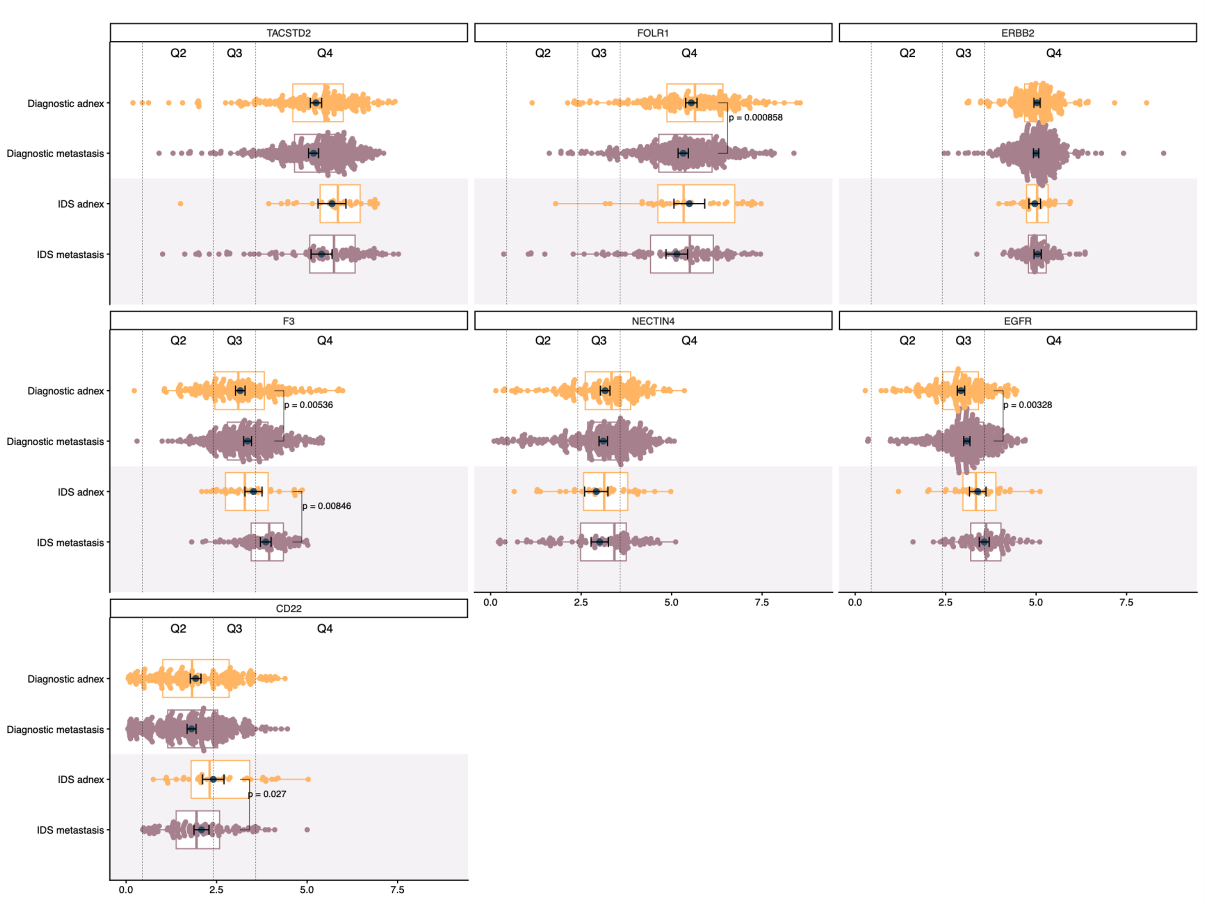


**Supplementary Figure 1. Effects of the varying tumor sites.** Input data as swarm- and boxplots, individually for each target. Results are divided into the studied treatment phases (diagnostic samples on white background and IDS samples on pale grey) and tumor sites (adnex samples with yellow and metastasic samples in taupe). Estimated marginal means, confidence intervals, and significant differences between the tumor sites are marked for each target. The quartiles from the expression landscape are illustrated with vertical dashed lines.

**Supplementary Figure 2. Target expression across diagnostic and IDS samples.**

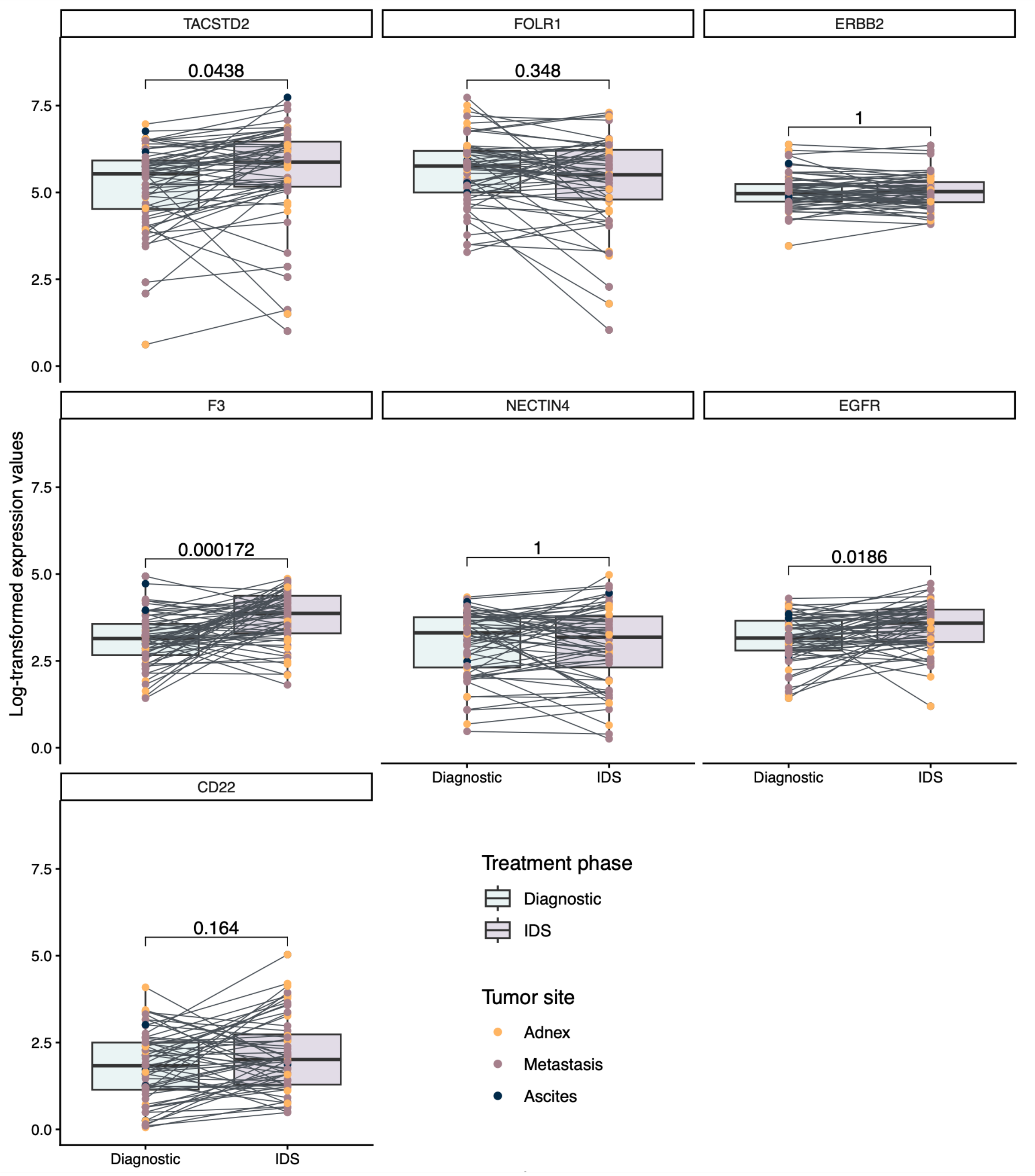


**Supplementary Figure 2. Target expression across diagnostic and IDS samples.** The figure illustrates the bulk RNA expression of patient matched diagnostic and IDS samples. Each sample is colored based on the collected tumor site, and each diagnostic–IDS pair relates to a line.

### Supplementary Figure 3. The concordance of intra-tumor heterogeneity and bulk RNA expression.

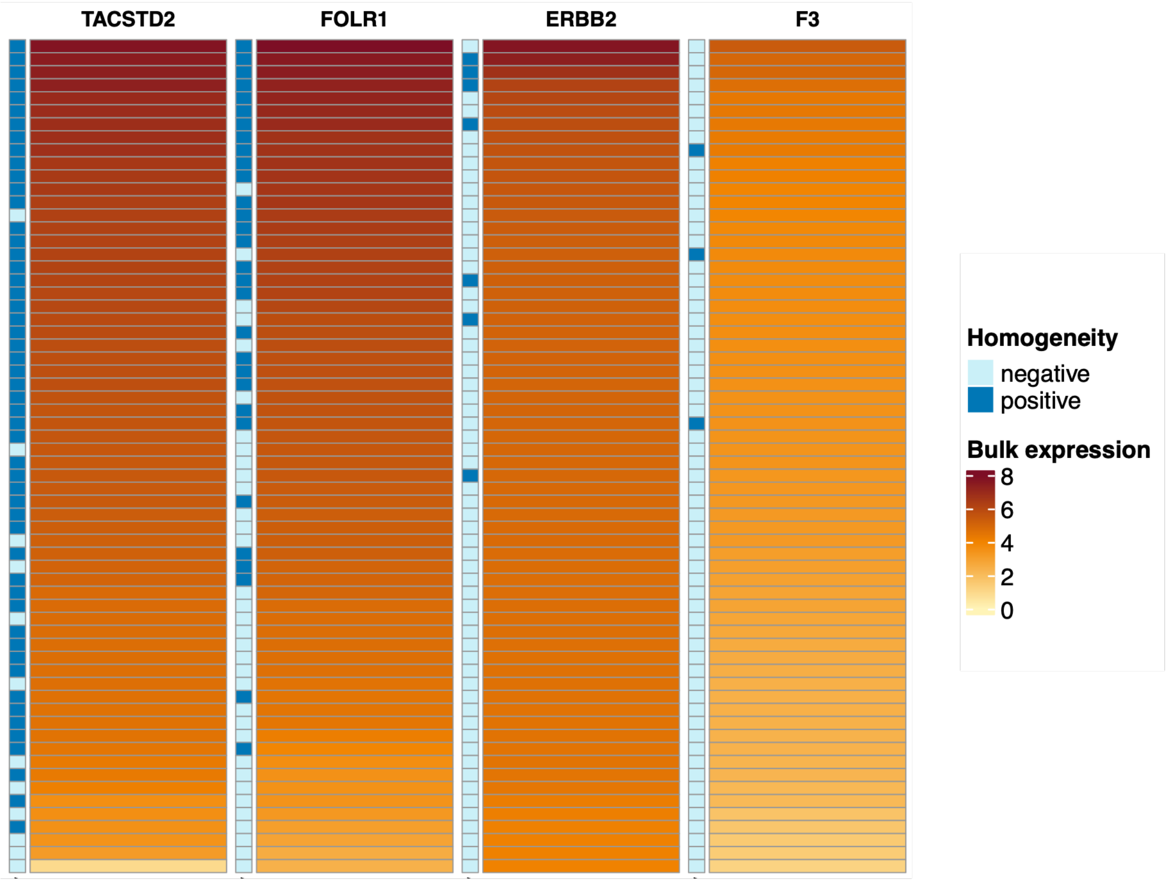


**Supplementary Figure 3. The concordance of intra-tumor heterogeneity and bulk RNA expression.** Bulk RNA expression compared to the intra-tumor heterogeneity from the matching scRNA-seq samples. Each of the genes is ordered based on descending bulk RNA expression, and the corresponding intra-tumor heterogeneity is marked in the annotation column.

**Supplementary Figure 4. Expression relationship between FOLR1 and TACSTD2.**

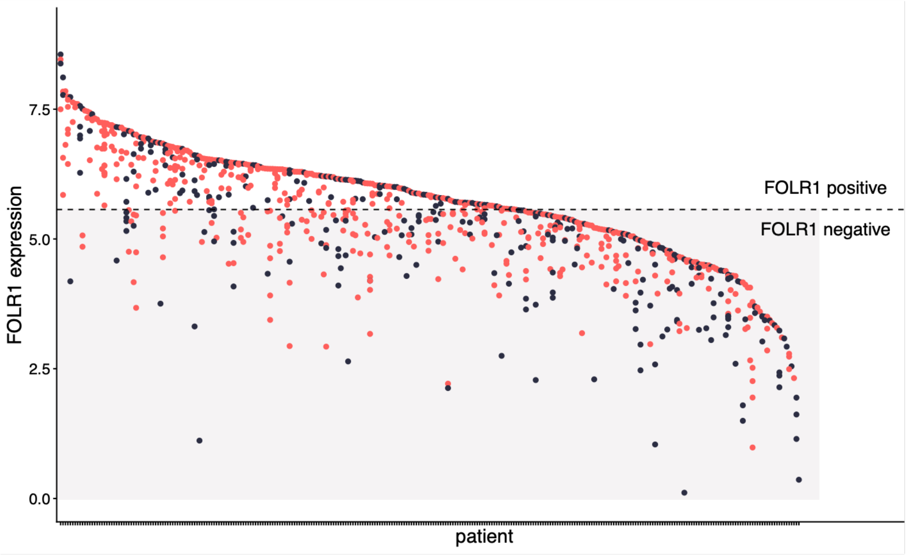


| | TACSTD2 positive | TACSTD2 negative |
|---|---|---|
| **FOLR1 positive** | 264 | 122 |
| **FOLR1 negative** | 286 | 195 |

**Supplementary Figure 4. Expression relationship between FOLR1 and TACSTD2.** FOLR1 and TACSTD2 expression across all samples. Points are ordered by FOLR1 expression and classified as intratumor positive (homogeneous) or negative (heterogeneous) using ROC-derived thresholds from matched bulk and intratumor heterogeneity analysis samples (Supplementary Figure S2); TACSTD2 positivity is indicated by color. Summary statistics are shown in the contingency table.
